# Evaluation of a Cook Islands Māori model of palliative care: a protocol

**DOI:** 10.64898/2026.02.11.26346011

**Authors:** Amy Henry, El-Shadan Tautolo, Josephine Herman, Jan Dewar, Teremoana Maua-Hodges, Ioana Mulipola

## Abstract

**Aim:** This research aims to evaluate the effectiveness, cultural appropriateness, and feasibility of the Cook Islands palliative care model *te vaerua kōpū tangata ora* within palliative care practice.

**Background:** Access to palliative and end of life care is a recognised human right, yet significant disparities persist for Pacific peoples in Aotearoa, New Zealand. While the understanding of different cultural perspectives has grown, in Aotearoa, there remain gaps in the delivery of culturally appropriate palliative care.

**Methodology:** This study will use a Cook Islands Tīvaevae research methodology to guide semi-structured interviews with 25-35 Cook Islands community members and 10 palliative care clinicians. This approach will support a rich, relational, and culturally grounded exploration of how a Cook Islands Māori palliative care model can be integrated into clinical practice.

**Discussion:** Recommendations to improve culturally responsive palliative care will be formulated in collaboration with community members and clinicians. The study will contribute to the limited body of knowledge on Pacific cultural understandings of palliative care and provide practical insights into applying an indigenous Pacific model within the palliative care system.

## Introduction

Research in palliative and end-of–life care that offers insight into the existing palliative care knowledge of diverse community groups is an area worthy of exploration. Development and adaptation of Pacific approaches in the field of palliative care, that are developed from Pacific paradigms are likely to deliver novel insights on palliative and end-of-life care. A model for Cook Islands Māori palliative care was created in the primary researcher’s PhD. The model - *Te vaerua kōpū tangata ora*, translates to the wellness of the family spirit. The model is a culturally responsive model of holistic palliative care designed specifically for Cook Islands Māori in New Zealand (NZ)(Henry, 2024). The model emphasises holistic, family driven care and considers the spiritual wellness of the family while centring the palliative family member, as they move through a spiritual transition. The model is inclusive of the role of the Tupuna (ancestors) and the importance of maintaining family harmony. The model was developed using grounded theory methodology alongside the Cook Islands research methodology, Tīvaevae. The model was formulated as a conceptual model through interviews with Cook Islands Māori caregivers, Cook Islands Māori clinicians and traditional Māori healers (n=28). The objective of this study is to test suitability of the model to the wider Cook Islands Māori community in Aotearoa, NZ and to determine the suitability of the model to the clinical setting.

## Background

The Cook Islands holds a unique place within the socio-political environment of Aotearoa, NZ. Cook Islands Māori are connected to NZ Māori through familial, cultural and historic ties. The Cook Islands is made up of 15 islands in the South Pacific. The Cook Islands were annexed to NZ in 1901 (Scott, 1991) and later became a self-governing nation in free associated with NZ in 1965(Smith, 2010). Due to the cultural and historical relationship between Māori of NZ and the Cook Islands, the Cook Islands were governed through the department of Native affairs, NZ. During this period many prominent NZ Māori politicians held roles in visiting and governing the Cook Islands, including Sir Apirana Ngata, (Gilson, 1980)Te Heuheu Tūkino (Tūkino, 1911)and Maui Pomare (Pomare, 1912). This historical relationship between the Cook Islands and NZ continues today.

Today, there is an estimated 94,176 Cook Islands Māori residing in NZ (Statistics, 2023) and around 27,000 Cook Islands Māori in Australia (Australian Bureau of Statistics, 2023), compared to 17,000 people residing in the Cook Islands (Cook Islands Statistics Office, 2025). The Cook Islands is part of the NZ realm, as an independent nation in free association with NZ. Part of being a realm nation includes Cook Islands Māori having unharboured rights to NZ citizenship and residency. Given the high percentage of Cook Islands Māori residing in NZ as a population, it is debated, a health system that is responsive to the needs of Cook Islands people are essential for the well-being of the Cook Islands people, both in NZ and in the homelands. Health models and policies which reflect the relationships between NZ and Pacific nations are important steps towards acknowledging our shared history and future within the realm of NZ.

### Pacific Peoples use of Palliative Care Services

The World Health Organization [WHO] (World Health Organization, 2021) describes palliative care as an ethical necessity that should be accessible to all and adapted to suit the needs of the local cultures. Community engagement is promoted as the primary strategy to meet the needs of local communities and cultures (World Health Organization, 2018). In line with the recommendations of the WHO, the current NZ Palliative Care Strategy (2001) states that all people in need of palliative care “should have access to quality palliative care services that are culturally appropriate” (Ministry of Health, 2001) additionally stating the need to understand the needs of Pacific cultures. However, currently there is limited research on the palliative care needs for Pacific peoples and even less research focusing on palliative care for Cook Islands Māori peoples in NZ.

There is limited data about palliative care use among Pacific peoples in Aotearoa NZ. However, available evidence suggests distinct patterns in how Pacific communities engage with palliative care reflecting cultural norms of caring for family members at home (Foliaki et al., 2020). Pacific peoples with cancer are more likely to die at home than NZ Europeans (Gurney et al., 2022). For example, earlier research reported 30% of Pacific peoples died at home (Palliative Care Council of New Zealand, 2011) while another found even higher rates, up to 42% compared to 36% of Māori and 27% of NZ Europeans, within one NZ city (Taylor et al., 2011). This pattern aligns with broader evidence showing that many Pacific peoples volunteer significant time to caring for a family or community members and providing for the community (Ministry of Pacific peoples, 2021). The survey found that over 20% of Pacific families regularly care for an elderly family member or community member, while over 10% regularly care for a member with an illness, and around 10% regularly care for a person with a disability (Ministry of Pacific peoples, 2021). Given these cultural norms and caregiving patterns, palliative care planning and policy needs to account for Pacific concepts of care, service and family responsibility to ensure equitable distribution of palliative care resources.

Differences in how Pacific people understand and conceptualise palliative and end of life, impacts on their use of palliative care services (Henry et al., 2025; Tuala, 2018). There are few resources tailored specifically for Pacific communities and there is a lack of understanding within Pacific communities of NZ-based palliative care services (Foliaki et al., 2020). In addition, there is a reported lack of resources in Pacific languages and lack of palliative care community engagement with Pacific communities (Foliaki et al., 2020; Henry, 2020).

### Methodology

The study will use the Tīvaevae methodology to study how the palliative care model (*Te vaerua kōpū tangata ora*) can be translated into practice. The Tīvaevae is a Cook Islands Māori research methodology designed by (Maua Hodges, 2018) and it has been used by numerous researchers of Cook Islands and non-Cook Islands descent. Over time the Tīvaevae methodology has been built upon, as researchers adapted the model to their research and developed the model into a methodology that can be applied and used to guide a wide range of research. Tīvaevae is the name given to a traditional Cook Islands quilt and is traditionally made by a group of women (Rongokea, 1992). Five key values of the Tīvaevae were developed by Te Ava (2011) which will be used to guide this study. These values are taokotai – collaboration; tu akangateitei – respect; uriuri – reciprocity; tu inangaro – relationships; and *akāri kite* – a shared vision (Te Ava & Rubie-Davies, 2011).

The Tīvaevae methodology is based on Cook Islands Māori traditions and paradigms. Like the creation of a Tīvaevae (quilt) it is created through working together as a group. A shared vision is described by Te Ava and Rubie-Davies (2011) as the objective of the methodology. The ontological underpinning of the Tīvaevae is embedded in *akāri kite* (a shared vision). It is within the relationships formed that that our realities, are tested against each other, created and shaped. It is for this reason that the model ‘*te vaerua kōpū tangata ora*, will be strengthened by the methodology.

## Study design

### Study aims and objectives

The aim of this research is to evaluate the effectiveness, cultural appropriateness, and feasibility of the Cook Islands palliative care model ‘*te vaerua kōpū tangata ora’* in palliative care. The objectives of the research:

- To examine the cultural acceptability by exploring community members perceptions of the model’s alignment with traditional beliefs and practices.
- To evaluate the feasibility by identifying barriers and facilitators in implementing the model within the existing healthcare systems.
- To compare clinicians’ perceived outcomes between the model and conventional palliative care approaches.
- To determine the transferability of the model to the wider Pacific community in NZ and abroad.

### Study design overview

The study is split into four phases, gap analysis, adaptation, feedback and review and implementation.

### *Phase one* - gap analysis

will review the cultural and clinical appropriateness and transferability to practice through qualitative focus groups with members of the Cook Islands Māori community and palliative care clinicians. This process involves deeply understanding and affirming the model’s cultural relevance, effectiveness, and appropriateness within the community it serves. The first phase of validation ensures the model is not only grounded in Indigenous knowledge systems but also practical, sustainable, and meaningful for the community, and clinicians, fostering trust and supporting its integration into healthcare practices.

### *Phase two* – adaptation

will adapt the model for use in the clinical setting, creating a practice-based framework for the implementation of the model into practice.

### Phase three – feedback and review

the model will be disseminated through the community focus groups for final feedback and review.

### *Phase four* – implementation

involves reviewing the application of the model, in practice, by interviewing clinicians over an eight-month period. Interviews will occur at 3-4 months and to 6-8 months after education on the model has been provided.

### Phase one: *Tāmoumou te Tīvaevae* (to secure)

Group interviews, three to four groups, the first set of interviews (*tāmoumou te Tīvaevae*) to secure the foundation of the Tīvaevae, will include community members (n 15-20). These interviews will introduce the current model ‘*te vaerua kōpū tangata ora’*. The model is currently a conceptual model, the purpose of these interviews is to ground the model through relational knowledge and lived experience. The interviews will be recorded and analysed in real time and post interviews.

The second set of interviews (*tuitui te Tīvaevae*-to sew), will present the model alongside the findings from the community to clinicians (n-10). This will mean the clinicians will be interviewed after the foundation has been set by the community. This method similar to Kaupapa Māori is to honour Cook Islands knowledge as the experts in creating a Tīvaevae.

The clinicians will then be interviewed on the clinical application of the model.

### Phase two: *Tuitui te Tīvaevae* (to sew): Amending the model

Creating the practical steps to apply the model in practice. In this phase the data collected from the previous interviews will be analysed and used to amend the model.

### Phase three: *‘Akamānea te Tīvaevae* (to decorate): community interviews

The model will be presented to the community (n=10-15) for final review (see Table 3). This will include a presentation of the model and a focus group review. Collaboration or working with the community is part of the creation process for making a Tīvaevae (Te Ava & Rubie-Davies, 2011), discovering shared objectives and leaning into the varied perspectives of others (Te Ava & Rubie-Davies, 2011). The model will be finalised and education resources created.

**Table 1:** *Tāmoumou te Tīvaevae* - community interviews. Focus: implementation of ‘te vaerua kōpū tangata ora’

|  |  |
| --- | --- |
| *Number of participants* | 15-20 community members |
| *Focus of interviews* | Spiritual needs of the family<br>Psycho-social needs of the family<br>Physical needs of the family<br>Practical needs of the community<br>Needs to be determined by the community |
| *Inclusion criteria* | Cook Islands heritage<br>Interest or experience in palliative care, aged care, end of life, chronic health care, primary health care |

**Table 2:** *Tuitui te Tīvaevae* (to sew)-clinician interviews. Focus: dependent on the needs identified by the community

|  |  |
| --- | --- |
| *Number of participants* | 10 clinicians (registered and/or unregistered) |
| *Inclusion criteria* | Currently working in palliative care generalist or specialist, community palliative care providers, hospice, primary health care, community NGO health care providers, aged care, chronic, long term health care<br>Clinicians defined as registered clinicians, eg. Nurses, doctors, social workers, non-registered clinicians eg health care workers, community liaison |
| *Focus of interviews* | Review what the model aims to achieve<br>Review practical clinical implementation of the model<br>Consider role of health care providers in implementation<br>Expected outcomes of the model<br>Plan for sustainability |

**Table 3:** ‘Akamānea te Tīvaevae. Focus: to refine and finalise the model

|  |  |
| --- | --- |
| *Number of participants* | 10-15 community members |
| *Focus of interviews* | Final review of the model by the community<br>Determine if the key components hold true<br>Discuss how the model will be implemented in practice<br>Gather feedback |
| *Inclusion criteria* | Involved in previous community interviews<br>Cook Islands heritage<br>Interest or experience in palliative care, aged care, end of life, chronic health care, primary health care |

**Table 4:**
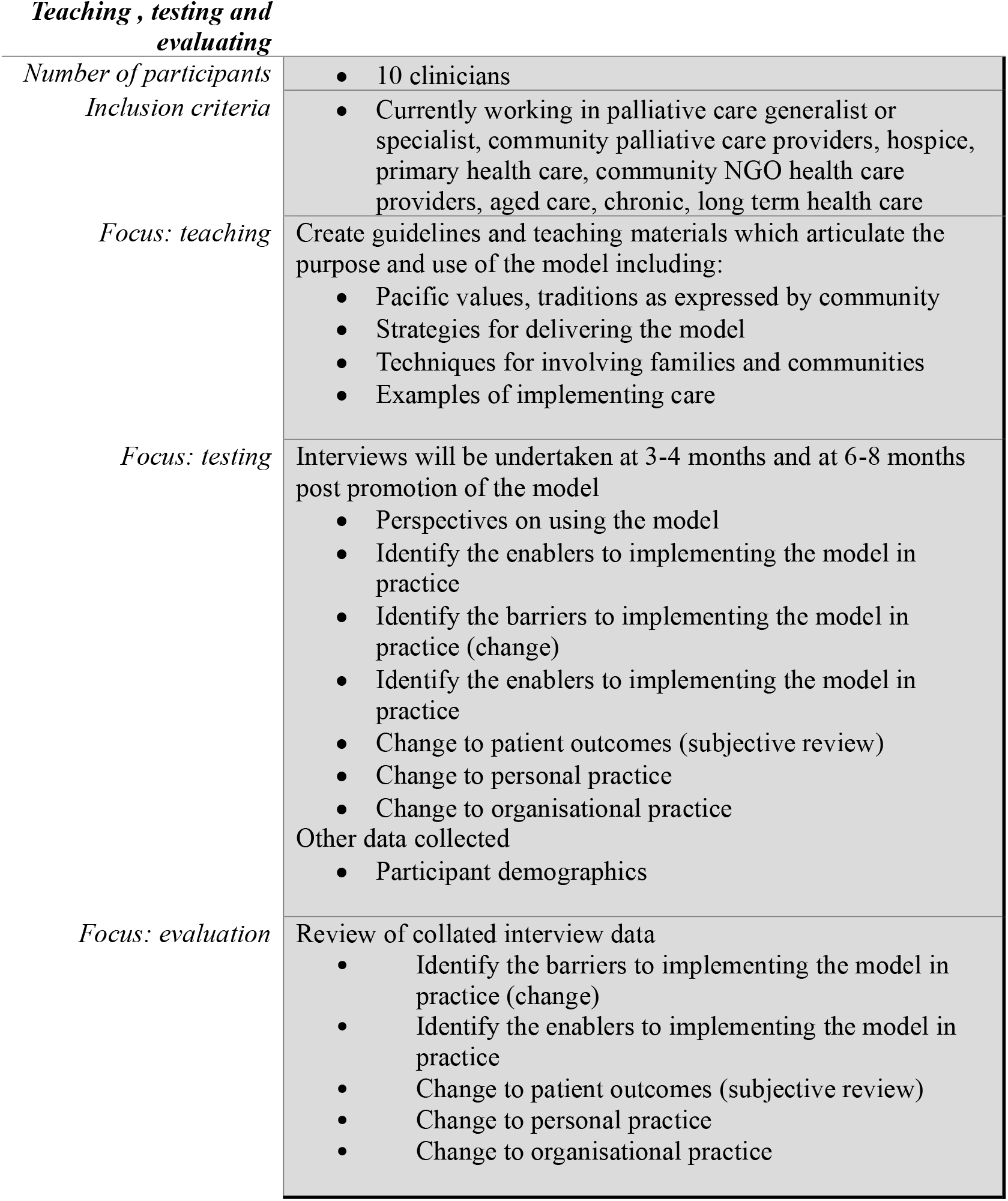
‘Akāiri’anga. Focus: to evaluate the model in practice

### Phase four: *‘Akāiri’anga* (to hang, to put in place)

Will include presenting the model to clinicians and community members. Specific teaching and engagement with clinicians to promote and facilitate adoption of the model.

## Recruitment

Phase one, three and four: Recruitment for these phases will involve using a purposive and snowball strategy for both groups. For in-person meetings, community members and clinicians will be recruited from Auckland, NZ, Auckland being home to the greatest number of Cook Islands Māori families.

### Community members

Community members will be recruited from community organisations through networking and advertising to the community.

Collaboration: work with and seek guidance from steering group members to design and implement recruitment process. Advertise study on Cook Islands platforms. Attend community events to personally explain the study and answer questions.

### Clinicians

will be recruited from hospice, community primary health care and NGO practices who have experience in the field of palliative care.

## Sample size

In this qualitative study, rich description and in-depth exploration of both community and clinician perspectives is sought. The sample size is therefore guided by the need to balance feasibility with meaningful depth of understanding. Consistent with the Tīvaevae methodology – which emphasises a collective and shared vision (Te Ava, 2011) - the sample size was determined with consideration of this concept. Data saturation will be reached when interviews yield no new insights, and no additional codes are generated during analysis. The previous study in which the model was developed (Henry, 2024) included 28 participants, providing a strong foundation of community knowledge. This study builds on that foundations to further develop and refine a shared vision.

## Data collection

Phases one, two and four: Data collection will use a semi-structured interview guide, which will be continually adapted as the interviews progress.

### Interviews

a semi-structured (*‘āpa’āpa* – half full) interview guide has been created to provide structure for consistency while remaining flexible and responsive to participants’ contributions. The guide is grounded in Cook Islands Māori concepts as developed by Te Ava Te Ava (2011) - including *tu inangaro* (relationships), *uriuri* (respect), and *akaāri kite* (shared vision), which serve as guiding principles for applying the Tīvaevae methodology. The layering concept of *‘akapapa’anga* (genealogy or the act of layering) is also incorporated as a foundational step. Powell (2013) describes *‘akapapa’anga* (Powell, 2013) as “*the relationality between individuals or cultural elements who are all part of an ever-growing whole or body”* (p.132) emphasising the interconnectedness that underpins the knowledge-building process.

The semi -structured guide will remain dynamic and adaptable, shifting as new insights emerge in a continuous circle of knowledge creation. This approach aligns with other iterative qualitative methodologies, such as grounded theory, by recognising participants as contributors to, and co-creators of, emerging knowledge. Conceptually, this process mirrors the making of a Tīvaevae – adding stiches to build a pattern, or unpicking areas where the pattern has not yet fully formed – reflecting the collective development of understanding.

## Data analysis

Although explicit data analysis methods are not described in the existing literature on the Tīvaevae methodology, a set of guiding principles spans the entire research process. These guidelines provide a flexible and open strategy that can be interpreted and adapted to suit the needs of individual studies. Maua Hodges (2018) demonstrates how the practical application of the Tīvaevae model can be applied across disciplines, providing a culturally grounded approach to conceptual development and synthesis. When applied to the coding of qualitative data, the Tīvaevae process offers a clear outline of the sequential steps – securing, stitching, and refining – that can inform a culturally aligned and systematic coding strategy.

There are three key stages in the coding and analysis of qualitative data that align closely with the stages of creating a tīvaevae, and these can be used to guide the analytical process. The creation stages include *tāmoumou* - to fasten or secure; *tuitui* - to sew; and *‘akamānea* - to decorate. As described by Maua Hodges (2018), *tāmoumou* te tīvaevae involves the pange (tīvaevae group) tacking the tīvaevae together and securing the initial patterns. This is followed by *tuitui te tīvaevae*, where each member of the tīvaevae group sews their assigned section, collectively checking and monitoring the emerging work. The final stage, *‘akamānea te tīvaevae involves tidying, refining and decorating* the tīvaevae to complete the piece (Maua Hodges, 2018).

From this interpretation of the Tīvaevae methodology, the following stages to data analysis will be applied. Interviews will be conducted in stages. The first stage, *tāmoumou* (to secure), will involve completing a set number of interviews, with the final number guided by (*piri’anga* – relationship, connection) the relationships that emerge between concepts during the interview process. Data analysis will occur concurrently with data collection, allowing for continual checking, refining, and validating of new information. Once key concepts have been developed through the stages of *tāmoumou*, the process of *tuitui* (to sew) will begin. In this stage, new interviews will be used to check and refine the emerging concepts for accuracy and coherence. The final stage, *‘akamānea* (to decorate), involves bringing together tthose who participated in the individual or family interviews for a group discussion to further shape, enhance, and “tidy” the Tīvaevae, ensuring that a shared and collectively affirmed vision has been created.

## Ethical considerations

Ethical approval was granted by AUT ethics committee (#25/105)

The ethical responsibilities of palliative care research in Pacific communities requires the researcher to maintain duty of care to the participants. Duty of care includes ensuring the research is based on Pacific paradigms and values. The Tīvaevae methodology closely aligns the research to Cook Islands Māori values and principles. This requires the research to be adaptable to meet the needs of the community as the community sees fit.

Research of palliative care raises unique ethical considerations and gaining ethical approval for research in palliative care studies has been documented as challenging in the literature (Gysels et al., 2013). Research in palliative care can be potentially triggering for participants and requires sensitivity by the researcher to monitor for signs of distress and end interviews if required. The primary investigator is a Cook Islands Māori and a clinically experienced registered nurse, who will be present at all interviews and will monitor for signs of distress and refer to follow-up care as required. Potential risks including distress from discussing sensitive issues will be monitored and a plan to minimise potential harm implemented (National Ethics Advisory Committee, 2019).

## Discussion

This study represents an innovative approach to collecting data on the cultural suitability of a Pacific palliative care model for the community and clinicians. The study addresses an area where very little is known, but new knowledge is evidently needed. As populations age and demands on palliative care services rise, addressing the unmet needs of underserved groups is become an increasingly urgent priority. Equally important is the engagement of community members to ensure that interventions are culturally responsive and meaningful for the intended community group. Engaging communities and working closely with palliative care clinicians will likely identify the strengths and barriers within the current palliative care system, and its ability to adapt to accommodate cultural models of care. Secondly, adapting the Tīvaevae for use in the palliative care setting aids in refining the methodology, to develop its theoretical foundations and demonstrate its practical application within the healthcare setting.

## Data Availability

Relevant deidentified research data from this study may be made available upon study completion and publication.

